# Are Remitters at Risk for Lower Food Security and Dietary Quality? An Exploratory Study of Mexican Immigrants in NYC

**DOI:** 10.1101/2022.12.20.22283288

**Authors:** Daniela Cruz-Salazar, Neil S. Hwang, Shirshendu Chatterjee, Kathryn P. Derose, Karen R. Flórez

## Abstract

Remittances confirm an immigrant’s continued social membership in their country of origin and they have been associated with improved nutritional outcomes among those who receive them. Yet, the relationship between remittances and nutrition outcomes of remitters is not well understood. We use data from 81 Mexican immigrants living in the Bronx, New York City (“NYC”), collected in 2019 to examine the relationship among remittances, gender, food security and dietary quality. After controlling for sociodemographic and immigration-related factors, we did not find a statistically significant (p<0.1) relationship between sending remittances and food insecurity; however, we did find that women remitters had higher odds than men remitters of having low dietary quality (p<0.064). We also found that a higher Body Mass Index (“BMI”) was associated with higher odds of experiencing low and very low food security (p<0.068). Further research with nationally representative data is needed to investigate the full extent of association between remittances and nutritional outcomes of remitters.

## Background

From 1990 to 2020, global immigration grew significantly. According to the United Nations, the total number of immigrants worldwide reached 281 million globally in 2020, representing 3.6% of the world’s population, up from 2.9% in 1990 [1]. The United States (“US”) continues to host the most number of immigrants (51 million, 15.5% of the US population), followed by Germany (16 million, 19.2% of Germany’s population) and Saudi Arabia (13 million, 37.3% of Saudi Arabia’s population) [2]. The countries where the largest number of international immigrants come from include India (18 million, 1.3% of India’s population), Mexico (11 million, 8.5% of Mexico’s population), and Russia (11 million, 8.4% of Russia’s population) [3]. Most immigrants currently living in the US come from Mexico, accounting for 24% of the total immigrant population, followed by India (6%), and China (5%) [4]. Mexican immigrants, in particular, tend to concentrate in a single country of destination. As of 2020, the US hosted 97% of all Mexicans living abroad [5].

The immigration patterns between Mexico and the US are a result of a complex set of geopolitical and economic factors between the two nations. Historically, US immigration policy has been crafted to source the labor needs of the American economy [6]. However, since 2016, there has been a decrease in net immigration to the US from Latin America, as the policy approach has shifted towards militarizing the US-Mexico border [7]. In addition, the Border Patrol and Immigration and Customs Enforcement have hired more agents, coinciding with increased budgets for apprehension and detention both along the border. Coupled with this shift in US immigration policy is the increased political unrest in some parts of Latin America that has effectively resulted in a humanitarian crisis. Chaos, suffering, and misery are routinely seen at the borders, meanwhile families and children continue to seek refuge [8].

The promise of economic stability in the US remains the driving engine for most immigrants coming from Mexico and elsewhere, as 77% of Hispanics believe they can get ahead with hard work compared to 62% of the general US public [9]. Remittances, defined as money transfers by immigrants to their relatives or other persons in their countries of origin [10], are an important aspect of immigrant’s life. Even with drops in income due to the COVID-19 pandemic, global remittances reached $650 billion in 2020 (representing 0.77% of the global Gross Domestic Product) [11]. The US is the top country for remittance outflows ($68 billion), and Mexico is the third largest recipient country for remittance inflows ($43 billion) [12]. Remittance flows into Mexico in 2020 were three times higher than Foreign Direct Investment, making them the largest source of financial flows into the country [13].

### Overview of Remittances with a Focus on Health

The impact that remittances have on economic growth for the countries that receive them (“destination countries”) has been widely researched. Empirical evidence suggests that remittances can contribute positively to economic growth in the destination countries by alleviating food insecurity and increasing savings and investment [14]. More specifically, a study focused on Middle East and North Africa countries found that remittances are used for investments such as human capital, housing, and land rather than household consumption, and that there is a positive relationship between economic growth and remittances [15]. Additionally, in South Asia, economic growth from remittances has been achieved through education and financial sector development [16]. A study of nine countries including Mexico found that remittances used for educational expenditures, energy use, and income significantly boosted economic growth over the long run [17].

In addition to investments, evidence suggests that food purchases are a major use of cash from remittances across 60 developing countries [18]. In rural Mexico, remittances received from the US reduced food insecurity at the household level [19]. In another study, households receiving international remittances in Nepal were found to be more food secure than those households that did not receive such remittances [20].

While there is extensive research on the nutritional impact of remittances on the people that receive them, the relationship between remittances and the nutrition status of *the people who send them* (“remitters”) is not well understood. Most related studies have focused on the motives immigrants may have to send money to their home countries [21-24] or on the impact remittances may have on the mental health of remitters [25-27]. To the best of our knowledge, only two studies have examined the food security levels of the remitter immigrants. One study from 2008 found that remitters were more likely to report hunger, but no meaningful associations were found for gender [28]. The other study from 2013 found that gender roles were an important indicator for the differences in remittance behavior among undocumented Mexicans in NYC, where men (n=301) remitted more than women (n=130), 94% compared to 65%; however, there was no link between gender differences and food insecurity among remitters [29]. Our study examines this link in detail.

### Research Question and Hypotheses

Our study addresses this gap in the literature by focusing on gender, remittances, and dietary outcomes of those who send them. This is done in several ways. First, for the measure of food security, we use the one defined by the US Department of Agriculture (“USDA”) since it has consistently been linked to multiple deleterious health conditions [30]. The second domain of our analysis includes dietary quality measures, since food insecurity is associated with lower dietary quality, including low consumption of nutrient-dense foods such as vegetables and fruits [31]. Third, our study focuses on Mexican immigrants living in the Bronx Borough of NYC (“the Bronx”), one of the largest immigration hubs in the US. The hypotheses we test in this paper relate to the subpopulation of Mexican Immigrants residing in the Bronx and include a) remitters are more likely to suffer from food insecurity compared to those who do not remit; b) remitters are more likely to have lower dietary quality compare to those who do not remit; and c) the effect is more pronounced for women than men.

## Methods

Data come from a study that explored in-depth the social networks, and dietary behaviors and outcomes of 81 Mexican immigrants recruited from a Catholic Church in the Bronx between January 2019 and June 2019. Inclusion criteria for the study included age (≥18 years of age) and ethnicity-nationality (self-identifying as Chicano, Mexican, or Mexican American). Trained bilingual research assistants obtained informed consent and collected data in their preferred language. All procedures are subjected to full review by the CUNY SPH Institutional Review Board, and all protocols were approved (protocol number 2018-1081).

### Measures

Our conceptual framework, research questions focused on remittances, gender, food security, and a decision tree analysis (shown in Figure 1) guided our decisions regarding variable selection and thresholds for constructing binary data.

**Figure 1.**
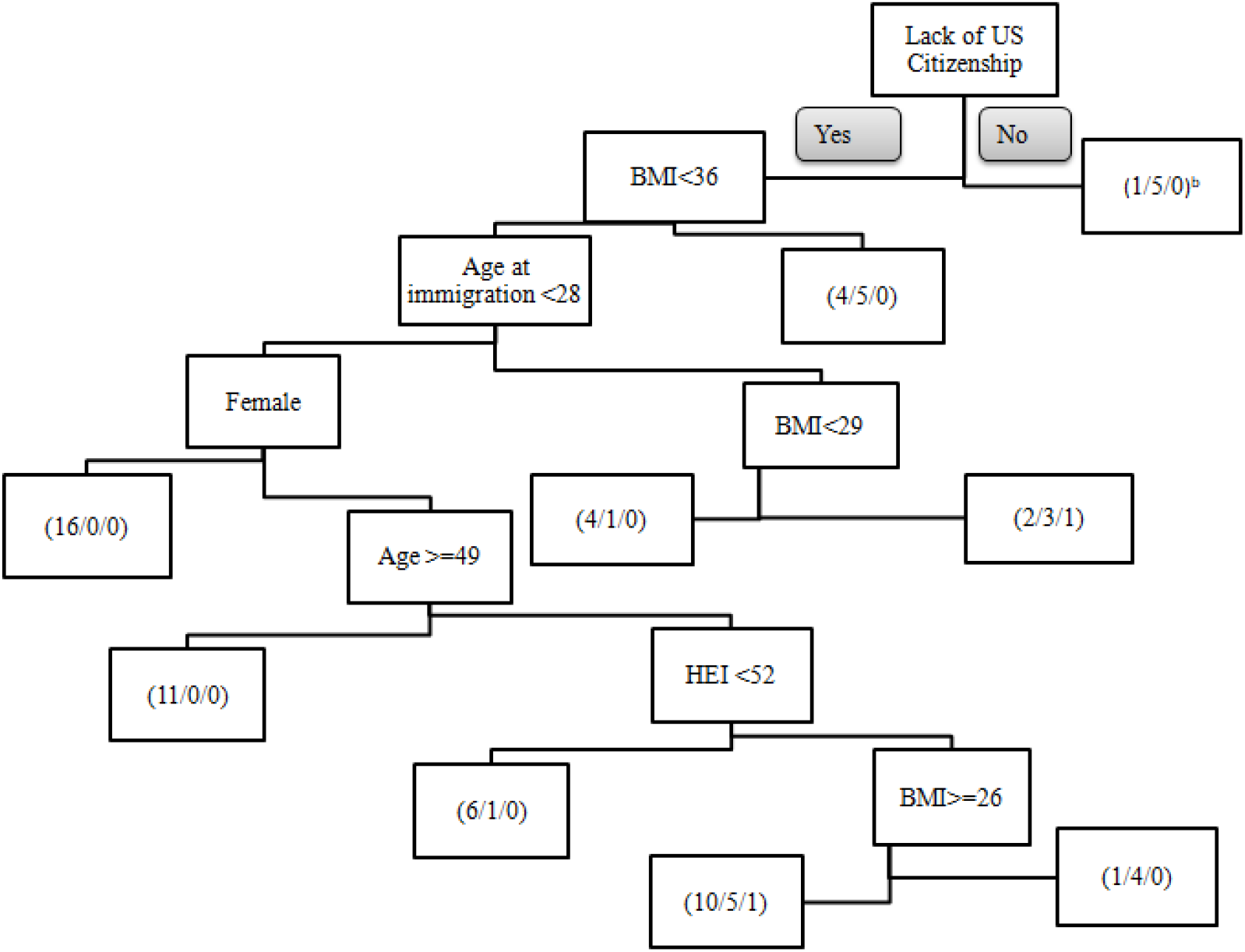
Decision tree with food security as the dependent variable for the analysis (N=81^a^) ^a^ Decision trees are robust to missing values since the counts in each node are based on the observed values only through that level in the tree. For this reason, N= 81 for our decision tree, but N=77 in our regression analysis. ^b^ Food security levels (High or marginal/ Low/ Very low)

### Dependent variables

#### Food security

Food security is defined as reliable access to sufficient and healthy food that one can afford [32]. The USDA define ranges of food security with the following labels: “high food security refers” to no reported indications of food access problems or limitations; “marginal food security” refers to one or two reported indications, such as anxiety over food sufficiency or a shortage of food at home, but little or no indication of changes in diet or food intake; “low food security” refers to reports of reduced quality, variety, or desirability of diet, but little or no indication of reduced food intake; and “very low food security” (formerly “food insecurity with hunger”) refers to multiple indications of disrupted eating patterns and reduced food intake [33]. Participants were asked the same questions as in the US Household Food Security Survey developed by the USDA in their preferred language [34]. The sum of affirmative responses to a specified set of items is referred to as the household’s raw score on the scale comprising those items [35]. Raw scores of 0 and 1 are classified as “high or marginal food security”; raw scores from 2 to 4 are classified as “low food security”; and raw scores from 5 to 6 are classified as “very low food security”. For our study, the food insecurity data was binarized into 1 and 0, with 1= denoting “low” and “very low food security” and 0= denoting “high” or “marginal food security”.

#### Dietary quality

The Healthy Eating Index (“HEI”) is a measurement tool of dietary quality that is scaled from 0 to 100, where 100 indicates completely alignment of a person’s diet with the 2015-2020 Dietary Guidelines of Americans [36]. There are 13 components, of which 9 measure adequacy (consumption of total fruit, whole fruit, total vegetables, greens and beans, whole grains, dairy, total protein foods, seafood and plant proteins, and fatty acids) and 4 measure foods to be consumed in moderation (refined grains, sodium, added sugars, and saturated fats). The score for adequacy components increases with increased consumption, whereas the score for moderation components increases with decreased consumption [37]. Participants’ dietary intake was assessed using a 24-hour dietary recall (Automated Self-Administered 24-hour Dietary Assessment Tool (“ASA24”) also available in Spanish [38]). The total food consumed by the participants was downloaded directly from the ASA24 algorithm. This algorithm calculates HEI scores by considering the 13 components previously mentioned. Continuous and binary data were used in this analysis. The binary data were obtained by assigning 1= to HEI scores of less than 52, and 0= to HEI scores greater than or equal to 52. This cut-off point was chosen based on the results of studies that included HEI and race/ethnicity in their analysis [39-41] and the decision tree results shown in Figure 1.

### Independent variables

#### Sends Remittances

Participants were asked whether they sent money transfers to family or friends in Mexico, and the variable was coded 1= yes and 0= no. This question was modified from the National Latino Asian American Survey (“NLAAS”), which is the only national survey to specifically ask about remittances. For more detailed information refer to Section 26. Finances (FN) in the questionnaire documents from the Disparities Research Unit’s NLAAS, also available in Spanish [42].

#### Gender

Participants were asked whether they identify themselves as female (coded as 1) or male (coded as 0). Gender identity is a multidimensional construct that could include non-binary terms [43]; however, such classifications are beyond the scope of our analysis.

### Control variables

For control variables, we identified the socioeconomic and demographic factors that have been reported in the literature to be associated with remittances, food security or dietary quality. These include:

#### Annual household income

Participants were asked to include only wages or stipends from employment (not including pensions, investments, interest, dividends, rent, social security, alimony or child support, unemployment insurance and armed forces or veteran’s allotment, and all income was measured before any taxes paid). Annual household income was classified into four categories as follows: 1= Less than $10,000, 2= between $10,000 and $29,990, 3= $30,000 to $49,990, and 4=$50,000 or higher.

#### Educational attainment

The participants were asked about their highest level of education completed. If a participant attended school outside the US, they were asked to mark the US equivalent. Responses were classified into four categories: 1= less than 11^th^ grade, 2= high school or general development education completed, 3= some college education without any degree, and 4= any college degree or higher.

#### Age

Participants were asked for the year in which they were born, and their age was calculated based on the date the interview took place.

#### Age at immigration

Participants were asked how old they were when they first came to the US. This question also came from NLAAS. Evidence suggests that more time spent in a foreign country may increase the risks of unfavorable dietary changes, especially when it comes to in the US and Canada [44]. Furthermore, in the US, immigrants have been found to be in better health than their US-born counterparts, but their health advantages consistently decrease as the duration of their US residence increases [45-46].

#### Lack of US citizenship

Modified from NLAAS, this question asked whether a participant was born as a US citizen or became a citizen through naturalization. There is evidence that suggests that households composed entirely of noncitizens are more likely to be food insecure compared to households composed of natural born citizens [47-49]. Benefits of obtaining US Citizenship include bringing family members to the US (allowing for reunification), becoming eligible for federal jobs (which may lead to a higher income), and accessing the Supplemental Nutrition Assistance Program benefits [50]. Lack of US citizenship in our study is classified as 1= “participants does not have US citizenship”, and 0= “participants does have US Citizenship”.

#### Acculturation

Acculturation scores were derived using the Marin’s Short Scale [51]. The scale primarily captures changes in language use such as the language one speaks at home, the language one speaks with friends, the language one reads in, and the language one thinks in. In total, twelve questions were asked. Scores of at most three indicate low acculturation, and greater than three indicates high acculturation. For our analysis acculturation was represented as a continuous variable with 2 decimal points.

#### Body Mass Index

Trained interviewers determined each participant’s ability to stand and found that all participants were able to stand on both feet. After this assessment, participants were asked to remove anything from their pockets before being asked to step on the scale, which was a SECA Robusta 813 digital scale [52] calibrated before each measure. The interviewers then used a portable height measuring board (SECA) to measure height. Height was recorded in meters and weight was recorded in pounds. Using the standard formula of the Centers for Disease Control and Prevention [53], a person’s weight in kilograms was divided by the squared high in meters. A BMI less than 25 means the person’s weight falls within the healthy weight range; a BMI between 25 and 29.9 means it falls within the overweight range; and a BMI of 30 or higher means the weight falls within the category of obesity [54]. For our analysis, continuous data was utilized, up to the first decimal point.

### Analysis

Any participant who did not know (n=1) or refused (n=3) to answer any given question was excluded from the data set, resulting in a final analytical sample of N=77. All descriptive and bivariate statistics, as well as binary logistic multivariate regression models, were computed using IBM SPSS Statistics Version 27 [55]. Two different models were built to test our hypotheses. The first model tests whether Mexican immigrant remitters in the Bronx are more likely to experience food insecurity than those who do not remit after accounting for control covariates. This hypothesis is tested using a binary logistic multivariate regression (see Model 1). The model also tests the hypothesis that females carry the weight of sending remittances more than men by adding the interaction term Remittances x Female.

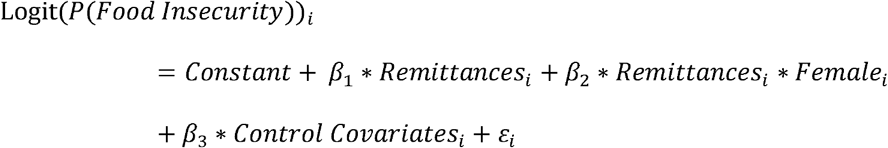

Logit refers to the natural log of the odds of a participant experiencing food insecurity. The adjusted odds ratio (“AOR”) is a measure of association between exposure (e.g., remittances) and outcome (e.g., food insecurity) [56], controlling for HEI, annual household income, education, gender, age, age at immigration, BMI, lack of US citizenship, and acculturation.

The second hypothesis tests the associations among Mexican immigrant remitters residing in the Bronx, and their dietary quality, compared to those who do not remit. This hypothesis is tested using the binary logistic multivariate regression (see Model 2). In addition, the interaction term Remittances x Female was included to test the significance of gender among remitters.

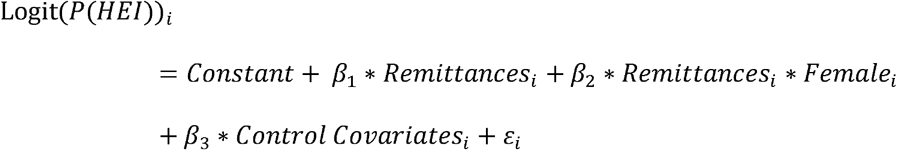

Logit (P(HEI)) refers to the natural log of the odds of a participant having a low HEI score (less than 52). The control covariates included food insecurity, annual household income, education, gender, age, age at immigration, BMI, lack of US citizenship, and acculturation.

We set the statistical significance level at 0.1 since this is a pilot study based on a relatively small sample with a robust set of control variables. This is consistent with various other studies published in public health with small community samples [57-61].

For the decision tree analysis, we used the R statistical software [62] for an exploratory analysis of the data. Decision trees are a useful tool for identifying the most informative covariates in modeling the dependent variable in an intuitive way. Decision trees are constructed with recursive binary partitions of the feature space [63].

Figure 1 shows a decision tree with food security as the dependent variable. The variables that best explain food security include HEI, gender, age, age at immigration, BMI and lack of US citizenship. For our dataset, the decision tree helps inform the split point for HEI (<52).

We briefly explain the output of the decision tree in Figure 1. For more detailed treatment of the methodology, refer to any standard text on machine learning, see e.g., [63-65]. Each box with a set of numbers is an end point. These end points describe how many participants following the tree’s conditions fall into each of the three categories of food security (High or marginal/ Low/ Very low). For example, the left most box that contains (16/0/0) means that for those who (1) lack US citizenship, (2) have a BMI of less than 36, (3) immigrated when they were younger than 28, and (4) are females, 16 of them experienced high or marginal food security, while none experienced low or very low food security.

## Results

Table 1 represents the descriptive statistics of our data set. It illustrates that in our sample, 37.7% of participants had low and very low food security (35.2% of women and 43.5% of men). In addition, HEI scores were roughly the same between men and women, with an average of 57.5. Regarding remittances, 70.1% of participants sent money transfers to Mexico, and a higher proportion of men did so (82.6%) than women (64.8%). Most of our sample (51.9%) had an annual household income less than $30,000, and education level of high school or less (81.8%). Our sample includes individuals aged between 20 and 68, with an average age of 43.5. The participants moved to the US when they were between 9 and 65 years old, and the mean age at immigration was 23.4. The average BMI among participants was 29.9, where women on an average were obese with a BMI of 30.2. The average acculturation score in our sample is 1.6 (reflecting low acculturation). Finally, only five participants in our sample had US Citizenship compared to 72 who did not.

**Table 1:** Descriptive characteristics of participants in the Bronx, NYC

|  | **Female (n=54)** | **Male (n=23)** | **Total (n=77)** |
| --- | --- | --- | --- |
| **Continuous Variables** | **Mean (25%, 75%)<sup>a</sup>** | **Mean (25%, 75%)** | **Mean (25%, 75%)** |
| Healthy Eating Index | 57.4 (51.0,65.4) | 57.5 (50.6, 67.6) | 57.5 (50.9,65.7) |
| Age | 44.4 (38.2, 51.0) | 41.4 (34.0, 54.0) | 43.5 (35.0, 51.0) |
| Age at immigration | 23.1 (17.0, 26.0) | 24.0 (17.5, 27.5) | 23.4 (17.0, 27.0) |
| Body Mass Index | 30.2 (26.0, 33.8) | 29.1 (26.4, 32.5) | 29.9 (26.2, 33.4) |
| Acculturation | 1.6 (1.2, 1.8) | 1.7 (1.3, 1.87) | 1.6 (1.3, 1.8) |
| **Categorical Variables** | **Count (%)** | **Count (%)** | **Count (%)** |
| Food Security |  |  |  |
| High/Marginal | 35 (64.8) | 13 (56.5) | 48 (62.3) |
| Low/Very Low | 19 (35.2) | 10 (43.5) | 29 (37.7) |
| Sends Remittances |  |  |  |
| Yes | 35 (64.8) | 19 (82.6) | 54 (70.1) |
| No | 19 (35.2) | 4 (17.4) | 23 (29.9) |
| Annual Household Income |  |  |  |
| Less than 30K | 33 (61.1) | 7 (30.4) | 40 (51.9) |
| 30K or higher | 21 (38.9) | 16 (69.6) | 37 (48.1) |
| Education |  |  |  |
| =<High school | 44 (81.5) | 19 (82.6) | 63 (81.8) |
| >=Some college | 10 (18.5) | 4 (17.4) | 14 (18.2) |
| Lacks US Citizenship |  |  |  |
| Yes | 51 (94.4) | 21 (91.3) | 72 (93.5) |
| No | 3 (5.6) | 2 (8.7) | 5 (6.5) |

<sup>a</sup> 25% denotes the 25<sup>th</sup> percentile and 75% denotes the 75<sup>th</sup> percentile.

Next, pairwise correlations were assessed to identify highly correlated covariates in the data set, as illustrated in Table 2. Gender and income have a low negative correlation of -0.308; in our sample, 61.1% of women had an annual household income of less than $30,000, compared to men at 30.4%. Similarly, lack of US citizenship and age had a low negative correlation (−0.369). Only two moderate positive correlations were observed. The first one was found for acculturation and education scores (0.556). The second was found with age at immigration and age (0.519).

**Table 2:**
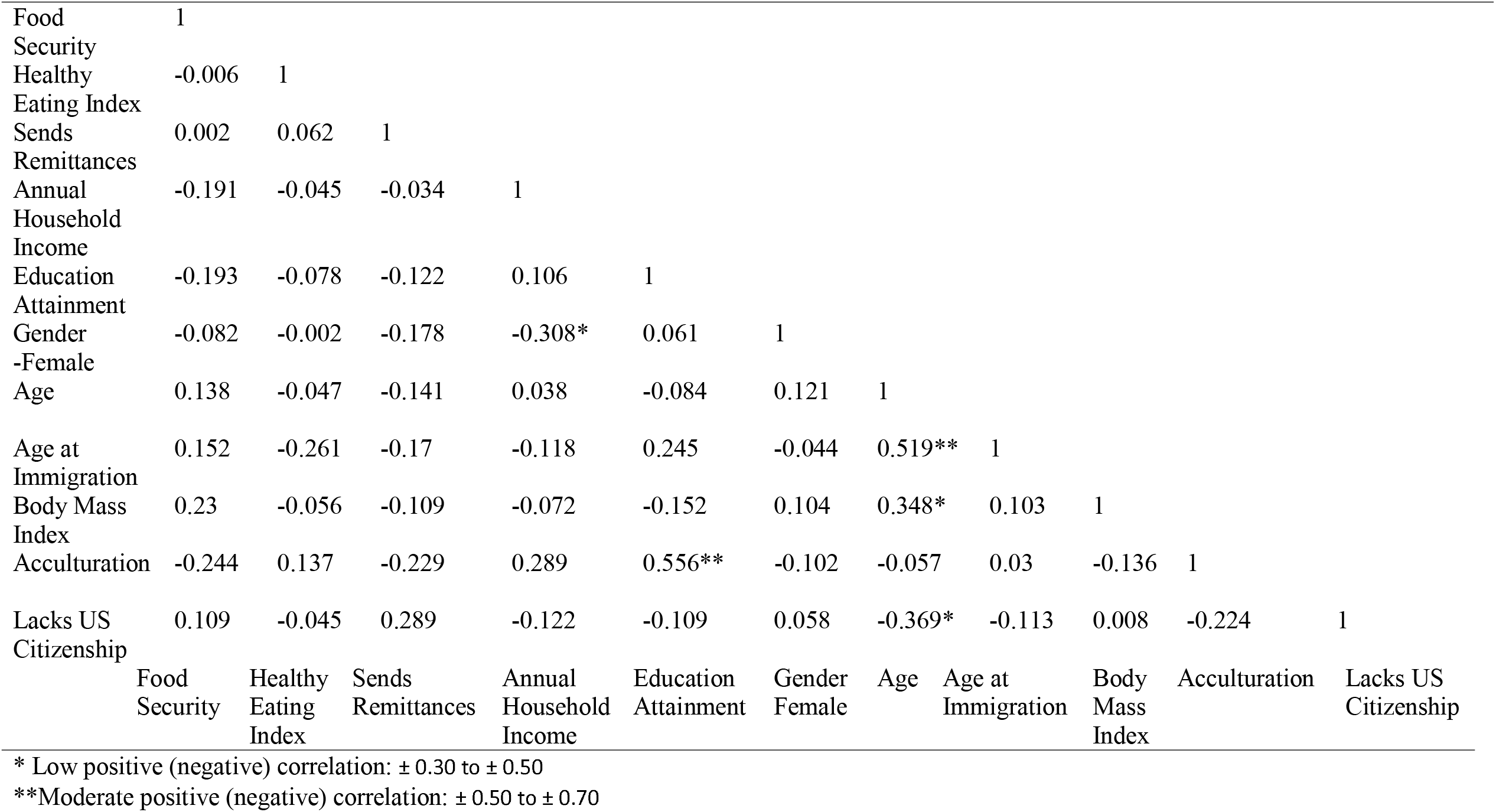
Pairwise correlations

|  |  |  |  |  |  |  |  |  |  |  |  |
| --- | --- | --- | --- | --- | --- | --- | --- | --- | --- | --- | --- |
| Food Security | 1 |  |  |  |  |  |  |  |  |  |  |
| Healthy Eating Index | -0.006 | 1 |  |  |  |  |  |  |  |  |  |
| Sends Remittances | 0.002 | 0.062 | 1 |  |  |  |  |  |  |  |  |
| Annual Household Income | -0.191 | -0.045 | -0.034 | 1 |  |  |  |  |  |  |  |
| Education Attainment | -0.193 | -0.078 | -0.122 | 0.106 | 1 |  |  |  |  |  |  |
| Gender -Female | -0.082 | -0.002 | -0.178 | -0.308* | 0.061 | 1 |  |  |  |  |  |
| Age | 0.138 | -0.047 | -0.141 | 0.038 | -0.084 | 0.121 | 1 |  |  |  |  |
| Age at Immigration | 0.152 | -0.261 | -0.17 | -0.118 | 0.245 | -0.044 | 0.519** | 1 |  |  |  |
| Body Mass Index | 0.23 | -0.056 | -0.109 | -0.072 | -0.152 | 0.104 | 0.348* | 0.103 | 1 |  |  |
| Acculturation | -0.244 | 0.137 | -0.229 | 0.289 | 0.556** | -0.102 | -0.057 | 0.03 | -0.136 | 1 |  |
| Lacks US Citizenship | 0.109 | -0.045 | 0.289 | -0.122 | -0.109 | 0.058 | -0.369* | -0.113 | 0.008 | -0.224 | 1 |
|  | Food Security | Healthy Eating Index | Sends Remittances | Annual Household Income | Education Attainment | Gender Female | Age | Age at Immigration | Body Mass Index | Acculturation | Lacks US Citizenship |

\* Low positive (negative) correlation: $\pm 0.30$ to $\pm 0.50$
\*\*Moderate positive (negative) correlation: $\pm 0.50$ to $\pm 0.70$

Table 3 shows the results for the binary logistic multivariate regression that tested the association between food insecurity and remittance senders as well as the gender interaction (see Model 1). This model estimated an AOR for BMI of 1.115 (p<0.068), meaning that higher BMI was associated with greater odds of having low or very low food security. In addition, Table 4 shows the analogous results for dietary quality (as measured by HEI) as the dependent variable (see Model 2). The results show an improvement in the p-value for remittances, but the result is not statistically significant at the 0.1 level. Women were less likely overall to have low HEI scores than men (AOR 0.012, p<0.070). However, the interaction term between remittances and gender is the highlight of our results. It shows that women who remitted had higher odds of having a low HEI score (OR=108.2, p<0.064) compared to men. Greater acculturation was significant and negatively associated with low HEI scores (OR=0.02, p<0.002), meaning more acculturated individuals were less likely to have low HEI scores. Finally, an older age at immigration was also positively associated with low HEI scores (OR=1.09, p<0.071).

**Table 3:** Logistic regression with food insecurity as the dependent variable^a^

| **Independent Variables** | **<u>Odds Ratio</u>** | **<u>95%CI</u>** |  | **<u>p-value</u>** |
| --- | --- | --- | --- | --- |
|  |  | **Lower** | **Upper** |  |
| Healthy Eating Index | 1.016 | 0.971 | 1.062 | 0.490 |
| Sends Remittances | 0.153 | 0.003 | 6.758 | 0.332 |
| Annual Household Income | 0.669 | 0.331 | 1.352 | 0.263 |
| Education Attainment | 0.667 | 0.330 | 1.348 | 0.259 |
| Gender- Female | 0.083 | 0.002 | 3.828 | 0.203 |
| Age | 1.012 | 0.945 | 1.084 | 0.727 |
| Age at Immigration | 1.037 | 0.952 | 1.130 | 0.403 |
| Body Mass Index | 1.115 | 0.992 | 1.254 | 0.068* |
| Acculturation | 0.477 | 0.106 | 2.143 | 0.334 |
| Lacks US Citizenship | 10.524 | 0.237 | 467.397 | 0.224 |
| Female x Remittances | 5.685 | 0.105 | 308.148 | 0.394 |
| Constant | 0.034 | - | - | 0.315 |

\*\*\*p<0.01, \*\*p<0.05, \*p<0.1
<sup>a</sup> 1=Low and very low food security and 0= High and marginal food security

**Table 4:** Logistic regression with low HEI as a dependent variable^a^

| **Independent Variables** | **<u>Odds Ratio</u>** | **<u>95%CI</u>** |  | **<u>p-value</u>** |
| --- | --- | --- | --- | --- |
|  |  | **Lower** | **Upper** |  |
| Food Security categories | 0.614 | 0.224 | 1.685 | 0.344 |
| Remittances | 0.026 | 0 | 2.936 | 0.130 |
| Annual household income | 1.643 | 0.759 | 3.558 | 0.208 |
| Education | 1.287 | 0.581 | 2.849 | 0.534 |
| Gender- Female | 0.012 | 0 | 1.444 | 0.070* |
| Age | 0.959 | 0.884 | 1.041 | 0.316 |
| Age at immigration | 1.090 | 0.993 | 1.196 | 0.071* |
| BMI | 1.061 | 0.936 | 1.203 | 0.352 |
| Acculturation | 0.020 | 0.002 | 0.229 | 0.002*** |
| Lack US Citizenship | 2.412 | 0.021 | 272.251 | 0.715 |
| Female x Remittances | 108.216 | 0.763 | 15343.254 | 0.064* |
| Constant | 141.232 | - | - | 0.205 |

\*\*\*p<0.01, \*\*p<0.05, \*p<0.1
<sup>a</sup> 1= low HEI (<52) and 0= high HEI (≥52)

## Discussion

Our cross-sectional analysis is a first step in elucidating the role of remittances on nutritional and health outcomes among immigrants. Our analysis focused on a community sample of Mexican immigrants living in the Bronx and suggests that women remitters within the sample were more likely to have a lower quality diet (HEI < 52) than men remitters. However, no significant relationships were found between food insecurity, gender, and remittances, as originally hypothesized. In contrast, we found that a higher BMI was associated with low and very low food security.

Remittances reflect an immigrant’s priorities, commitments, and personal relationships. In addition, remittances are an integral part of the lived experience of many immigrants, especially of those of Mexican descent [66]. Remittances confirm an immigrant’s continued social membership in their country of origin [67]. Remittances not only depend on the network left behind in the country of origin but also on the strength of the network in the immigrant’s current life [68]. Remittances are highly influenced within the context of one’s family; they can represent contractual patterns (a sense of obligation) among family members, or they can be ingrained from social norms rather than conscious decisions made by the immigrant [69]. They can also exert negative pressure on the individual, to the point where senders may skip or cut down their meals. Though our data did not provide evidence that remittances are associated with food insecurity, it does suggest that dietary quality could be negatively affected especially for women.

The most common motivations to remit are altruism, exchange, insurance, loan repayments, and investments [70-72]. Altruistic remittances are mostly sent to an immigrant’s primary family in the country of origin to improve their financial circumstances. Immigrants care deeply about those they left behind, and thus the emotions associated to remit are typically strong [73]. Altruistic remittances aim to improve the recipients’ income, consumption, and standard of living, despite negatively affecting the sender’s standard of living [74].

This could explain the results that Mexican immigrant remitters who are women are more likely to have a lower dietary quality than men. There is evidence to suggest that, in the case of women, they are, and are expected to be, more altruistic than men [75]. For example, in a study using a dictator game in which the participants (playing the role of a dictator) receive an endowment and then decides to what extent they want to split this endowment with another [76], it was shown that women chose to split their endowments more often than men [77]. Similarly, a similar study focused on adolescents showed that girls tended to be more altruistic than boys [78]. Another study of Amazon Mechanical Turk crowdworkers in the US found that not only were women more altruistic but they were also expected to be so by both genders [79]. A meta-analysis of 22 experimental studies found evidence suggesting that women are expected to be altruistic, and that the more a woman describes herself as independent or dominant compared to warm or tender, the less was her altruism [80]. In other words, the more a woman describes herself using male characteristics, the lower her altruistic scores became.

In addition to altruism, other characteristics unique to women may help explain our results. Immigrant women may be doubly disadvantaged. First, the disadvantages may arise from the discrimination they face as women and immigrants [81-82]. One way these disadvantages manifest themselves is via income inequalities. The pay gap between non-immigrant men and immigrant women in high income countries is estimated at 20.9%, which is greater than the aggregate gender pay gap of 16.2% [83]. In our analysis, we control for annual household income; however, due to the categorical nature of our data on annual household income, it could be that gender income disparities related to the double disadvantages women immigrants face explain our results.

In addition to gender, our results also indicate a relationship between the age at which participants in our sample first arrived in the US with dietary quality, after accounting for control variables. Our results suggest that a higher value for age at immigration is associated with higher odds of having a low HEI score (< 52). Our results also reveal an association between the acculturation score of an immigrant in our sample with dietary quality, after accounting for control variables. A higher level of acculturation is associated with lower odds of having low scores of HEI. Several factors could explain these results. Immigration comes in hand with acculturation, which often refers to changes in values, beliefs, attitudes, and behaviors [84]. High acculturation can also result in better economic outcomes, and there is evidence to suggest this may be associated with a greater consumption of vegetables for more acculturated immigrants [85-86]. However, opposite to our results, higher acculturation has also been linked empirically in some studies to poor food choices among immigrants [87], especially for women [88]. This could be because the more acculturated an immigrant becomes, the more integrated she will likely be into the standard American diet [89].

Furthermore, our analysis suggests that immigrants in our sample who have a higher BMI are more likely to experience low and very low food security. This result is supported by the existing literature. Food insecurity has been shown to be associated with poor diets and malnutrition [90-91]. High rates of obesity are more likely in low-income communities that have poor food environments or that exist in food desserts [92-93]. In addition, a study that reviewed 34 articles found that key barriers for healthy eating among young adults in Western countries include relatively low costs of processed foods, lack of time, knowledge and skills to plan, shop, and cook healthy foods, and the abundant presence of unhealthy foods in their environment [94].

Our study had several limitations that should be considered in the context of our results. First, the primary focus of the original study was not on the impact of remittances on health. As a result, our analysis was limited by the coarse granularity of our data set such as the absence of precise data on the amount or proportion of annual household income sent by immigrants to their country of origin; the frequency with which remittances are sent or the motives behind such money transfers. In addition, the exploratory nature of the study allowed for data collection from one community in the Bronx using non-probabilistic methods, which precludes our ability to generalize to the larger Mexican American population in the US. The small sample also contributed to lower precision for point estimates for our model parameters than what was needed for the significance level of 0.05. Finally, the observational nature of our study does not allow us to make any causal claims about the relationship between remittances and food security or dietary quality.

In summary, the findings of our study suggest that although female remitters might not be skipping their meals (i.e., experiencing acute food insecurity), they may still be compensating for the economic pressures by consuming a diet that is lower in quality. Acculturation and age at immigration are important factors for dietary quality as well. Yet for those who are experiencing acute forms of food insecurity, irrespective of gender and remittances, weight may be the relevant public health issue as indicated by the observation that most of the participants in our data set were either overweight or obese. Future work using a larger dataset could elucidate these important questions.

### Conclusion & New Contribution to the Literature

As mentioned in the introduction, most existing literature on remittances relates to their impacts on the people that receive them. Research on the effect remittances on food insecurity and the dietary quality of senders is scarce. Our analysis attempts to increase awareness of important topic, make initial findings that are meaningful as an initial step, and illustrate that more research is needed with nationally representative data.

## Data Availability

All data produced in the present study are available upon reasonable request to the authors

## References

[1] Our World in Data: Total number of international immigrants. https://ourworldindata.org/explorers/migration?tab=chart&facet=none&Metric=Number+of+international+immigrants&Period=Total&Sub-metric=Total&country=USA∼DEU∼FRA∼GBR∼SYR∼TUR∼YEM∼IND∼CAN∼OWID_WRL. Accessed 30 Oct 2022.

[2] United Nations Department of Economic and Social Affairs: International Migration 2020 Highlights. https://www.un.org/en/desa/international-migration-2020-highlights Accessed 30 Oct 2022.

[3] United Nations Department of Economic and Social Affairs: International Migration 2020 Highlights. https://www.un.org/en/desa/international-migration-2020-highlights. Accessed 30 Oct 2022.

[4] American Immigration Council: Immigrants in the United States, 2021. https://www.americanimmigrationcouncil.org/research/immigrants-in-the-united-states. Accessed 30 Oct 2022.

[5] American Immigration Council: Immigrants in the United States, 2021. https://www.americanimmigrationcouncil.org/research/immigrants-in-the-united-states. Accessed 30 Oct 2022.

[6] Portes A: Bifurcated immigration and the end of compassion. Ethnic and Racial Studies, 2020; 43:1, 2–17. DOI: 10.1080/01419870.2019.1667515

[7] Watson T, Thomson K: The Decline in U.S. Net Migration. EconoFact, 2022. https://econofact.org/the-decline-in-u-s-net-migration. Accessed 30 Oct 2022.

[8] Massey DS: The Real Crisis at the Mexico-U.S. Border: A Humanitarian and Not an Immigration Emergency. Sociological Forum 2020; 35: 787–805.

[9] Hugo-Lopez M, Gonzalez-Barrera A, Krogstad JM: Latinos are more likely to believe in the American dream, but most say it is hard to achieve. Pew Research Center, 2018. https://www.pewresearch.org/fact-tank/2018/09/11/latinos-are-more-likely-to-believe-in-the-american-dream-but-most-say-it-is-hard-to-achieve/. Accessed 30 Oct 2022.

[10] International Organization for Migration (IOM): Gender, Migration and Remittances. https://www.iom.int/sites/g/files/tmzbdl486/files/about-iom/Gender-migration-remittances-infosheet.pdf. Accessed 30 Oct 2022.

[11] The World Bank: GDP (Current US$) 1960–2201. https://data.worldbank.org/indicator/NY.GDP.MKTP.CD. Accessed 30 Oct 2022.

[12] Migration Data Portal: Remittances. https://www.migrationdataportal.org/themes/remittances. Accessed 30 Oct 2022.

[13] Ong R, Young D: Defying Predictions, Remittance Flows Remain Strong During COVID-19 Crisis. The World Bank, 2021. https://www.worldbank.org/en/news/press-release/2021/05/12/defying-predictions-remittance-flows-remain-strong-during-covid-19-crisis. Accessed 30 Oct 2022.

[14] Meyer D, Shera A: The impact of Remittances on Economic growth: An econometric model. EconomiA 2017; 18:2; 147–155.

[15] Ben-Mim S, Ben-Ali, MS: Through Which Channels Can Remittances Spur Economic Growth in MENA Countries? Economics 2012; 6:1; 2012–33.

[16] Cooray A: The Impact of Migrant Remittances on Economic Growth: Evidence from South Asia. Review of International Economics 2012; 20: 985–998.

[17] Zaman S, Wang Z, Zaman Qu: Exploring the relationship between remittances received, education expenditures, energy use, income, poverty, and economic growth: fresh empirical evidence in the context of selected remittances receiving countries. Environ Sci Pollut Res, 2021; 28; 17865–17877

[18] Crush J, Caesar M: Introduction: Cultivating the Migration-Food Security Nexus. International Migration 2017, 55: 10–17.

[19] Mora-Rivera J, Gameren EV: The impact of remittances on food insecurity: Evidence from Mexico, World Development 2021; 140, 105349.

[20] Regmi M, Paudel KP: Food security in a remittance based economy. Food Sec. 2017; 9, 831–848.

[21] Holst E, Schäfer A, Schrooten M: Remittances and Gender: Theoretical Considerations and Empirical Evidence. IZA DP 2011; 5472.

[22] Abrego L, LaRossa R: Economic Well-Being in Salvadoran Transnational Families: How Gender Affects Remittance Practices. Journal of Marriage and Family 2009; 71:4; 1070– 1085.

[23] Carling J: Scripting Remittances: Making Sense of Money Transfers in Transnational Relationships. International Migration Review 2014; 48(1_suppl), 218–262.

[24] Piracha M, Saraogi A: Motivations for Remittances: Evidence from Moldova. IZA DP. 2011; No. 5467.

[25] Amoyaw JA, Abada T: Does helping them benefit me? Examining the emotional cost and benefit of immigrants’ pecuniary remittance behaviour in Canada. Social science & medicine, 2016; 153; 182–192.

[26] Balasca C: Countervailing Effects? Remittance Sending and the Physical and Mental Health of Migrants. Thesis Graduate School of the Ohio State University, 2019. https://etd.ohiolink.edu/apexprod/rws_etd/send_file/send?accession=osu1575466424352253&disposition=inline. Accessed 30 Oct 2022.

[27] Ambugo EA, Yahirun JJ: Remittances and risk of major depressive episode and sadness among new legal immigrants to the United States, Demographic Research 2016; l34:8; 243−258.

[28] Hadley C, Galea S, Nandi V, Nandi A, Lopez G, Strongarone S, Ompad D: Hunger and health among undocumented Mexican migrants in a US urban area. Public Health Nutrition 2008; 11:2, 151–158.

[29] López GP: The Health of Undocumented Mexicans in New York City. Chicana/o Latina/o Law Review 2013; 32:1.

[30] Seligman HK, Berkowitz SA: Aligning Programs and Policies to Support Food Security and Public Health Goals in the United States. Annual Review of Public Health, 2019; 40:1, 319–337.

[31] Hanson KL, Connor LM: Food insecurity and dietary quality in US adults and children: a systematic review. The American Journal of Clinical Nutrition, 2014; 100:2, 684– 692.

[32] Oxford Learner’s Dictionary: Food Security Definition. Oxford University Press https://www.oxfordlearnersdictionaries.com/us/definition/english/food-security. Accessed 30 Oct 2022.

[33] US Department of Agriculture (USDA): Definitions of Food Security. Economic Research Service. https://www.ers.usda.gov/topics/food-nutrition-assistance/food-security-in-the-u-s/definitions-of-food-security/ Accessed 30 Oct, 2022.

[34] US Department of Agriculture (USDA): US Household Food Security Survey Module—Spanish: Three-Stage Design, With Screeners. Economic Research Service, 2013. https://www.ers.usda.gov/media/8285/hh2012spanish.pdf Accessed 30 Oct, 2022.

[35] US Department of Agriculture (USDA): US Household Module Food Security Survey Module three-stage design, with screeners. Economic Research Service, 2012. https://www.ers.usda.gov/media/8271/hh2012.pdf. Accessed 30 Oct 2022.

[36] US Department of Agriculture (USDA): Healthy Eating Index. https://www.fns.usda.gov/healthy-eating-index-hei. Accessed 30 Oct 2022.

[37] US Department of Agriculture (USDA): HEI-20151 Components and Scoring Standards. https://fns-prod.azureedge.us/sites/default/files/healthy_eating_index/HEI-2015%20Components%20and%20Scoring%20Standards_2.pdf. Accessed 30 Oct 2022

[38] National Cancer Institute: Automated Self-Administered 24-Hour (ASA24®) Dietary Assessment Tool. Division of Cancer Control & Population Science. https://epi.grants.cancer.gov/asa24/. Accessed 30 Oct 2022.

[39] Ma Y, Weng X, Gao X, Winkels R, Cufee Y, Gupta S, Wang L: Healthy Eating Index Scores Differ by Race/Ethnicity but Not Hypertension Awareness Status among US Adults with Hypertension: Findings from the 2011-2018 National Health and Nutrition Examination Survey. Journal of the Academy of Nutrition and Dietetics, 2022; 122:5; 1000–1012.

[40] Arandia G, Sotres-Alvarez D, Siega-Riz AM, Arredondo EM, Carnethon MR, Delamater AM, Gallo LC, Isasi CR, Marchante AN, Pritchard D, Van Horn L, Perreira KM: Associations between acculturation, ethnic identity, and diet quality among U.S. Hispanic/Latino Youth: Findings from the HCHS/SOL Youth Study. Appetite, 2018; 129; 25–36.

[41] Overcash F, Reicks M: Diet Quality and Eating Practices among Hispanic/Latino Men and Women: NHANES 2011-2016. Int. J. Environ. Res. Public Health, 2021; 18:3: 1302.

[42] The Mass General Research Institute: Questionnaire documents from the Disparities Research Unit’s National Latino and Asian American Study (NLAAS). https://www.massgeneral.org/mongan-institute/centers/dru/research/past/nlaas-docs. Accessed 30 Oct 2022.

[43] Current Measures of Sexual Orientation and Gender Identity in Federal Surveys. Federal Interagency Working Group on Improving Measurement of Sexual Orientation and Gender Identity in Federal Surveys, 2016. https://s3.amazonaws.com/sitesusa/wp-content/uploads/sites/242/2014/04/current_measures_20160812.pdf. Accessed 30 Oct 2022.

[44] Popovic-Lipovac A, Strasser B: A Review on Changes in Food Habits Among Immigrant Women and Implications for Health. J Immigrant Minority Health, 2015; 17, 582– 590.

[45] Frisbie WP, Cho Y, Hummer RA: Immigration and the health of Asian and Pacific Islander adults in the United States. American journal of epidemiology, 2001; 153:4; 372–380.

[46] Singh GK, Miller BA: Health, life expectancy, and mortality patterns among immigrant populations in the United States. Canadian journal of public health = Revue canadienne de sante publique, 2004; 95:3; I14–I21.

[47] Thomson RB: Food Insecurity in the U.S.: Does Citizenship and Birthplace Matter? The Journal of Public and Professional Sociology, 2022; 14:1; Article 1.

[48] Haro-Ramos AY, Bacong AM: Prevalence and risk factors of food insecurity among Californians during the COVID-19 pandemic: Disparities by immigration status and ethnicity. Preventive medicine, 2022; 164:107268. https://doi.org/10.1016/j.ypmed.2022.107268

[49] Kalil A, Chen JH: Mothers’ citizenship status and household food insecurity among low-income children of immigrants. New directions for child and adolescent development, 2008;121; 43–62. https://doi.org/10.1002/cd.222

[50] US Citizenship and Migrations Services (USCIS): What Are the Benefits and Responsibilities of Citizenship? A Guide to Naturalization, Ch 2. https://www.uscis.gov/sites/default/files/document/guides/chapter2.pdf. Accessed 30 Oct 2022.

[51] Marin G, Sabogal F, Marin BV, Otero-Sabogal R, Perez-Stable E: Development of a Short Acculturation Scale for Hispanics. Hispanic Journal of Behavioral Sciences, 1987; 9:2; 183–205.

[52] Seca Precision for Health: seca 813. https://www.seca.com/en_us/products/all-products/product-details/seca813.html Accessed 17 Dec 2022.

[53] Centers for Disease Control and Prevention (CDC): Adult BMI Calculator.https://www.cdc.gov/healthyweight/assessing/bmi/adult_bmi/english_bmi_calculator/bmi_calculator.html Accessed 17 Dec 2022.

[54] Centers for Disease Control and Prevention (CDC): Defining Adult Overweight & Obesity. https://www.cdc.gov/obesity/basics/adult-defining.html. Accessed 30 Oct 2022.

[55] IBM SPSS Statistics https://www.ibm.com/us-en?ar=1 Accessed 8th December 2022.

[56] Szumilas M: Explaining odds ratios. Journal of the Canadian Academy of Child and Adolescent Psychiatry = Journal de l’Academie canadienne de psychiatrie de l’enfant et de l’adolescent 2010; 19:3; 227–229.

[57] Yang Z, Paudel KP, Wen X, Sun S, Wang Y: Food Safety Risk Information-Seeking Intention of WeChat Users in China. Int. J. Environ. Res. Public Health, 2020; 17:2376. https://doi.org/10.3390/ijerph17072376

[58] Buman MP, Winkler EAW, Kurka JM, Hekler EB, Baldwin CM, Owen N, Ainsworth BE, Healy GN, Gardiner PA: Reallocating Time to Sleep, Sedentary Behaviors, or Active Behaviors: Associations With Cardiovascular Disease Risk Biomarkers, NHANES 2005– 2006, American Journal of Epidemiology, 2014; 179: 3; 323–334. https://doi.org/10.1093/aje/kwt292

[59] Rowley KG, Daniel M, Skinner K, Skinner M, White GA, O’Dea K: Effectiveness of a community-directed ‘healthy lifestyle’ program in a remote Australian Aboriginal community. Australian and New Zealand Journal of Public Health, 2000; 24: 136–144. https://doi.org/10.1111/j.1467-842X.2000.tb00133.x

[60] Wu S, Pan C, Yao L, Wu X: The Impact of the Urban Built Environment on the Play Behavior of Children with ASD. Int. J. Environ. Res. Public Health, 2022; 19:22; 14752. https://doi.org/10.3390/ijerph192214752

[61] Palesch YY: Some common misperceptions about P values. Stroke, 2014; 45:12; e244–e246. https://doi.org/10.1161/STROKEAHA.114.006138

[62] R-Project: What is R? https://www.r-project.org/about.html. Accessed 30 Oct 2022.

[63] Hastie T, Tibshirani R, Friedman J: The Elements of Statistical Learning -Data Mining, Inference, and Prediction. Califronia: Springer Series in Statistics, Second Edition; 2008. https://hastie.su.domains/Papers/ESLII.pdf. Accessed 30 Oct 2022.

[64] Mitchell, TM: Machine learning. V1: 9. New York: McGraw-hill, 1997. ISBN: 0070428077

[65] Mohri M, Rostamizadeh A, Talwalkar A: Foundations of machine learning. MIT press, Second Edition, 2018.

[66] Conway D, Cohen JH: Consequences of Migration and Remittances for Mexican Transnational Communities. Economic Geography, 1998; 74:1, 26–44.

[67] Carling J: Scripting Remittances: Making Sense of Money Transfers in Transnational Relationships. International Migration Review 2014; 48:1; 218–262.

[68] Holst E, Schäfer A, Schrooten M: Remittances and Gender: Theoretical Considerations and Empirical Evidence. IZA DP, 2011: No. 5472.

[69] Vanwey LK: Altruistic and Contractual Remittances between Male and Female Migrants and Households in Rural Thailand. Demography 2014; 41:4; 739–756.

[70] Yang D: Migrant Remittances. Journal of Economic Perspectives 2011; 25:3; 129–52.

[71] Piracha M, Saraogi A: Motivations for Remittances: Evidence from Moldova. IZA DP, 2011; No. 5467.

[72] Carling J: Scripting Remittances: Making Sense of Money Transfers in Transnational Relationships. International Migration Review 2014; 48:1; 218–262.

[73] Rapoport D, Docquier F: The Economics of Migrants’ Remittances. IZA DP, 2005; No.153.

[74] Vanwey LK: Altruistic and Contractual Remittances between Male and Female Migrants and Households in Rural Thailand. Demography 2014; 41:4; 739–756.

[75] Brañas-Garza P, Capraro V, Rascón-Ramírez E: Gender differences in altruism on Mechanical Turk: Expectations and actual behaviour, Economics Letters 2018; 170, 19–23.

[76] Leder J, Schütz A: Dictator Game. In: Zeigler-Hill, V., Shackelford, T. (eds) Encyclopedia of Personality and Individual Differences. Springer, Cham 2018.

[77] Cadsby CB, Servátka M, Song F: Gender and generosity: does degree of anonymity or group gender composition matter? Experimental Economics, 2010; 13, 299–308.

[78] Dreber A, von Essen E, Ranehill E: Gender and competition in adolescence: task matters. Exp Econ, 2014; 17, 154–172.

[79] Brañas-Garza P, Capraro V, Rascón-Ramírez E: Gender differences in altruism on Mechanical Turk: Expectations and actual behaviour, Economics Letters 2018; 170, 19–23.

[80] Rand DG, Brescoll VL, Everett JA, Capraro V, Barcelo H: Social heuristics and social roles: Intuition favors altruism for women but not for men. Journal of experimental psychology 2016; 145:4; 389–396.

[81] Adanu RMK, Johnson TRB: Migration and women’s health. International journal of gynecology and obstetrics, 2009; 106:2; 179–181.

[82] Lopez M: Skilled Immigrant Women in the US and the Double Earnings Penalty. Feminist Economics, 2012; 18:1, 99–134.

[83] Amo-Agyei S: The migrant pay gap: Understanding wage differences between migrants and nationals. International Labour Organization, 2020. ISBN 978-92-2-032371-7.

[84] Thomson MD, Hoffman-Goetz L: Defining and measuring acculturation: A systematic review of public health studies with Hispanic populations in the United States. Social Science & Medicine, 2009; 69:I7; 983–991.

[85] López EB, Yamashita T: Acculturation, Income and Vegetable Consumption Behaviors Among Latino Adults in the US: A Mediation Analysis with the Bootstrapping Technique. J Immigrant Minority Health, 207; 19, 155–161. https://doi.org/10.1007/s10903-015-0306-x

[86] Espinosa de Los Monteros K, Gallo LC, Elder JP, Talavera GA: Individual and area-based indicators of acculturation and the metabolic syndrome among low-income Mexican American women living in a border region. American journal of public health,2008; 98:11, 979–1986. https://doi.org/10.2105/AJPH.2008.141903

[87] Yoshida Y, Scribner R, Chen L, Broyles S, Philippi S, Tseng TS: Role of Age and Acculturation in Diet Quality Among Mexican Americans - Findings From the National Health and Nutrition Examination Survey, 1999-2012. Preventing chronic disease, 2017; 14, 170004.

[88] Popovic-Lipovac A, Strasser B: A review on changes in food habits among immigrant women and implications for health. Journal of immigrant and minority health, 2015; 17:2; 582–590.

[89] Pérez-Escamilla R: Acculturation, nutrition, and health disparities in Latinos. The American journal of clinical nutrition, 2011; 93:5, 1163S–7S. https://doi.org/10.3945/ajcn.110.003467

[90] Smith L, BarnettY López-Sánchez G F, Shin JI, Jacob L, Butler L, Cao C, Yang L, Schuch F, Tully M, Koyanagi A: Food insecurity (hunger) and fast-food consumption among 180164 adolescents aged 12-15 years from sixty-eight countries. The British journal of nutrition, 2022; 127:3; 470–477. https://doi.org/10.1017/S0007114521001173

[91] Keenan GS, Christiansen P, Hardman CA: Household Food Insecurity, Diet Quality, and Obesity: An Explanatory Model. Obesity Silver Spring Md, 2021; 29:1; 143–149. https://doi.org/10.1002/oby.23033

[92] Chen D, Jaenicke EC, Volpe RJ: Food Environments and Obesity: Household Diet Expenditure Versus Food Deserts. American journal of public health, 2016; 106:5; 881–888. https://doi.org/10.2105/AJPH.2016.303048

[93] Cheung HC, Shen A, Oo S, Tilahun H, Cohen MJ, Berkowitz SA: Food Insecurity and Body Mass Index: A Longitudinal Mixed Methods Study, Chelsea, Massachusetts, 2009–2013. Prev Chronic Dis, 2015;12:150001. DOI: http://dx.doi.org/10.5888/pcd12.150001

[94] Munt AE, Partridge SR, Allman-Farinelli M: The barriers and enablers of healthy eating among young adults: a missing piece of the obesity puzzle: A scoping review. Obesity reviews: an official journal of the International Association for the Study of Obesity, 2017; 18:1; 1–17. https://doi.org/10.1111/obr.12472

